# ELECTRONIC-BASED HEALTH PROMOTION MODELS ON THE USE OF POST PLACENTAL IUD BIRTH CONTROL: A SYSTEMATIC REVIEW AND META-ANALYSIS

**DOI:** 10.1101/2023.03.12.23286202

**Authors:** Henri Sulistiyanto, Soetrisno, A.A Subiyanto, Retno Setyowati

## Abstract

Low levels of IUD use, especially post-placental IUDs, are associated with bias and lack of information because contraceptive counseling is limited, for example, through conversations with doctors, written materials, or audiovisual techniques. This study aimed to identify electronic (video) interventions using post-placental IUD contraception through systematic review and meta-analysis. Based on those keywords, some results were obtained: 509 from Google Scholar, 33 from PubMed, 14 from Science Direct, 88 from Sage Journal, 50 from Springer Links, 20 from Scopus, and 178 from ProQuest. In addition, the recording was carried out on the bibliography of review articles, 12 articles were obtained for systematic review, and five meta-analyses were carried out. The heterogeneity test shows a value ρ greater than 0.05, which is ρ =0.88 with I^2^ = 0%, meaning that the variation between studies is homogeneous. The Random Effects Model yielded a pooled odds ratio of 1.07 (95& CI 0.26-4.39). The value ρ = 0.93 so that there was no difference between the treatment and control groups, and the data were homogeneous, an odd ratio of 1.07 was obtained, it was found that video use could increase 1.07 times the knowledge, skills, and decision-making of using post placental IUD compared to those who did not use video.

## INTRODUCTION

Worldwide, nearly half of the pregnancies are unwanted, with estimates ranging from 13% to 82%. Socio-demographic factors related to unwanted pregnancy include age, parity, education, and economic status. In addition to limited access to health services, barriers to contraceptive use include concerns about the safety or effectiveness of various methods of contraception and lack of knowledge or access to information about contraception (Dewart et al., 2019). Many interventions designed to prevent unwanted pregnancies have focused on patient education and contraceptive promotion to influence knowledge of contraception and perceived barriers to contraceptive use (Oulman et al., 2015).

Preventing unwanted pregnancies through effective contraceptive use is critical to a women’s health, but choosing between different contraceptive methods can be challenging, and opportunities for adequate discussion during regular consultations are often limited. Controlling fertility and being satisfied with the method of contraception chosen is very important for the health and well-being of women, but unwanted pregnancies remain common and expensive for individuals and health services. About 40% of pregnancies globally are estimated to be unplanned (Sedgh et al., 2014).

An RCT that integrates family planning into AIDS facilities shows a considerable effect on contraceptive use but a negligible impact on IUD use compared to injectable or implant use (Grossman et al., 2013). An RCT in Zambia directed at HIV couples found that video exposure to long-term contraceptive use effectively increased long-term contraceptive use. Among couples who did not use the method at first, 6% chose IUDs, 14% implants, 45% injections, and 33% oral contraceptives. Among those who already used contraception, 29% switched, mainly to long-term contraception (R. Stephenson et al., 2011).

Promotion concerning the use of IUDs is currently still low. Some respondents accept that implants are more straightforward to promote than IUDs and are favored by some institutions for various reasons so that the provider and client factors are involved in promoting the use of IUDs. Providers tend to like implants because they are not as complex as IUDs. The fact that implants are much more difficult to remove than IUDs is not explained to women. For women, implants are less intrusive than IUDs and, as a relatively new method, less appealing to misunderstandings and concerns (Cleland et al., 2017).

Research investigating contraceptive counseling is still limited, for example, through conversations with doctors, written materials, or audiovisual techniques. Some studies have assessed counseling for postpartum contraception but lost the opportunity for patients to initiate long-term reversible contraceptive methods or permanent contraceptives because their provision during labor periods is time-sensitive (Lopez et al., 2015). Previous research has shown that video-based contraceptive education can result in increased absorption. Others show increased knowledge and satisfaction, especially when paired with physician counseling (Davidson et al., 2015).

Common modalities for providing contraceptive education include communication with healthcare providers, written materials, audiovisual materials, and computer applications. With the increasing availability of mobile technology, using electronic platforms to deliver health education presents a unique opportunity for intervention. It can facilitate access to essential health information, especially in resource-limited areas. The patient’s report of eligibility and willingness to use electronic modalities for contraceptive education is generally favorable (McHenry et al., 2019).

Many interventions are designed to prevent unwanted pregnancies because they focus on patient education and contraceptive promotion to influence knowledge of contraception and perceived barriers to use (Xiong et al., 2021). Common modalities for providing contraceptive education include conversations with healthcare providers, written materials, audiovisual materials, and computer applications. With the increasing availability of mobile technology capabilities, electronic platforms for delivering health education present a unique opportunity for interventions to be disseminated in various settings (Prah, 2017). It can facilitate access to essential health information, especially in resource-limited areas. The patient’s report of eligibility and willingness to use electronic modalities for contraceptive education is generally favorable. However, a more formal evaluation of the effectiveness of the intervention delivered via electronic format is required (Modugu et al., 2018).

The challenge of carrying out promotions on less familiar and well-known methods, such as post-placental IUDs, is very severe. Providing comprehensive, well-balanced, and high-quality advice, information, and education necessitates more targeted and well-designed promotional approaches. Counseling of sufficient quality allows women to make the best decisions. Post-placental IUD insertion can be increased if antenatal counseling about birth control techniques is of higher quality. This systematic review and meta-analysis are intended to assess the effectiveness of electronics (video) as a tool for providing post-placental IUD contraceptive education. We searched for electronic interventions (videos) on post-placental IUD contraception.

## METHODS

In this study, data collection was conducted by systematically collecting articles in English scientific articles through the database of PubMed, Science Direct, Sage Journal, Springer Link, Scopus, ProQuest, and Google scholars using the keywords “video promotion model” AND “postpartum IUD” “health promotion model” OR video AND “postpartum IUD,” video OR m-health AND postpartum IUD. Three people conduct an abstract review to identify articles matching the research criteria. The article to be analyzed is selected if at least two reviewers agree with the case criteria.

Inclusion Criteria: (1) articles are limited to the last five years (2017-2022); (2) are original research articles; and (3) scientific research articles with all research designs. Exclusion Criteria: (1) the title is irrelevant; (2) no full text is available; (3) language other than English; (4) the abstract is irrelevant; (5) the deletion of scientific articles.

Based on those keywords, some results were obtained: 509 from Google Scholar, 33 from PubMed, 14 from Science Direct, 88 from Sage Journal, 50 from Springer Links, 20 from Scopus, and 178 from ProQuest. In addition, the recording was carried out on the bibliography of review articles, 12 articles were obtained for systematic review, and five meta-analyses were carried out. Systematic review and meta-analysis adhered to the PRISMA (Preferred Reporting Items for Systematic Reviews and Meta-Analysis) protocol for selecting articles to be analyzed, described in Figure 1.

**Figure 1.**
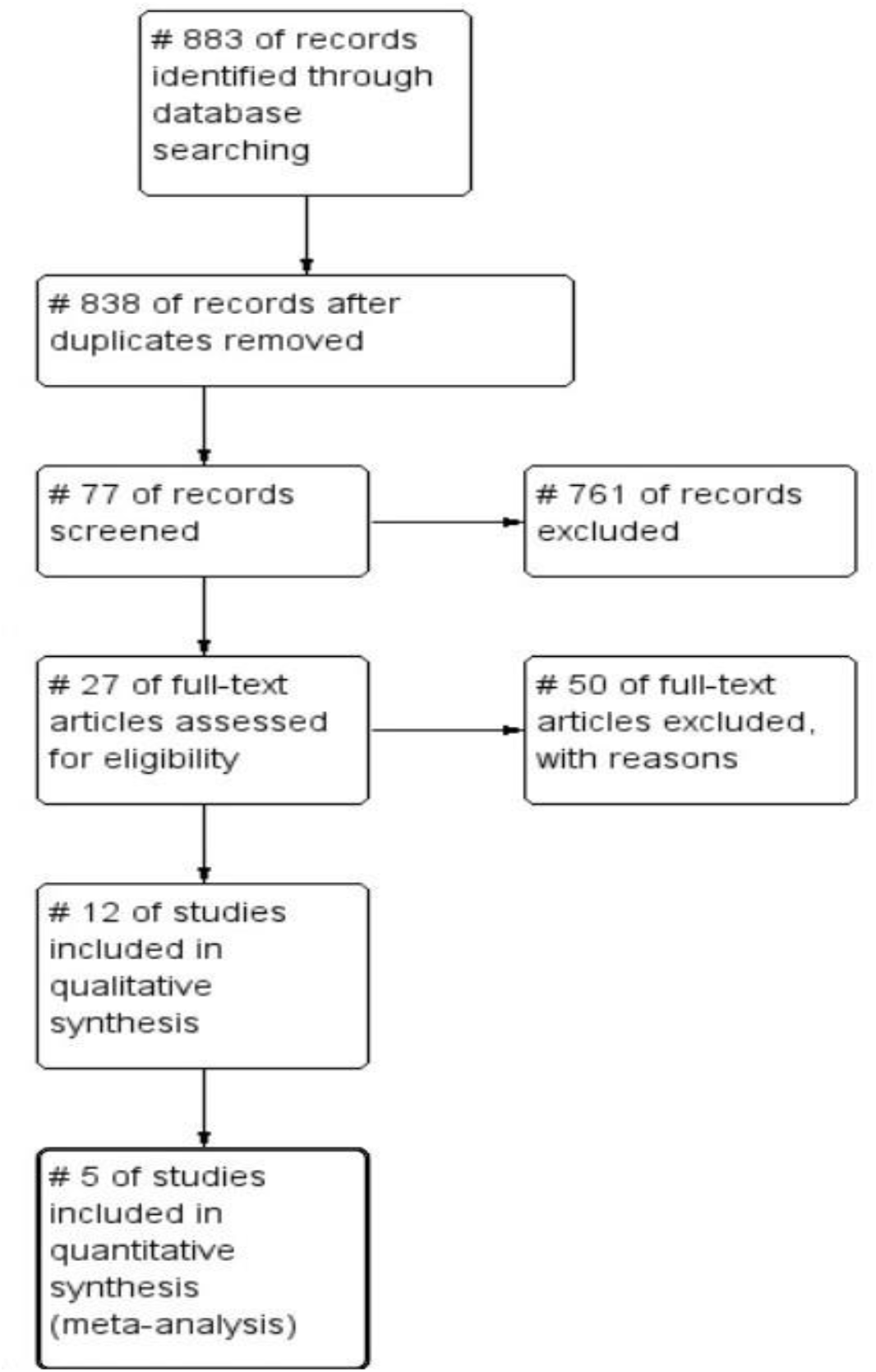
Systematic review protocol flowchart based on PRISMA (Preferred Reporting Items for Systematic Reviews and Meta-Analysis)

**Figure 2.**
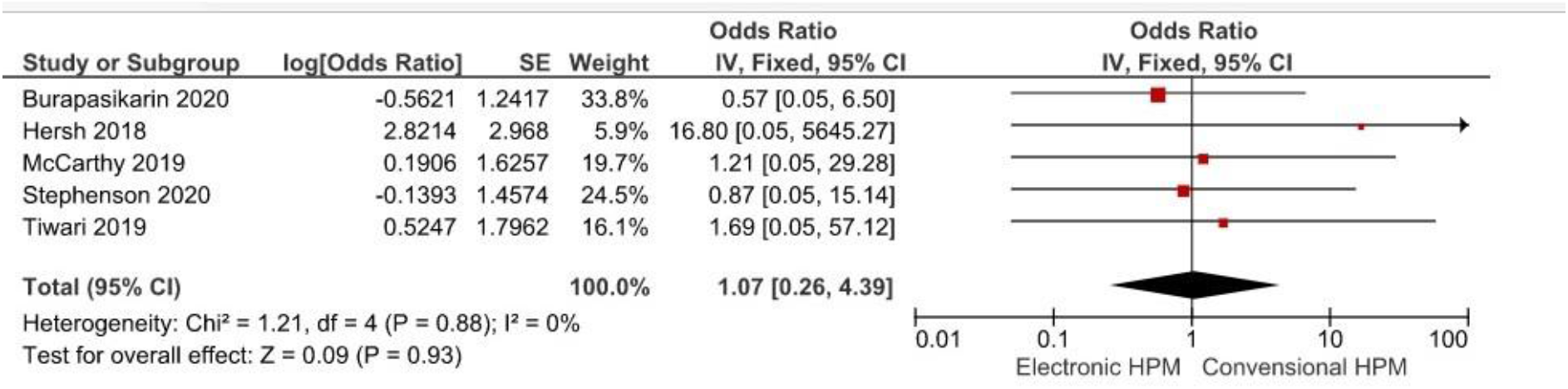
Forest Plot Diagram

Data analysis was conducted to obtain the pooled odds ratio, a combined odds ratio value from research using a fixed effect model using Haenszel’s Mantel method, and a random effect model. Using software version 5.3 of Review Manager, data were evaluated. The heterogeneity test is performed to establish the incorporation model for the meta-analysis. The statistical test I2 was performed to measure heterogeneity among several percentage-based research impact estimates. The Cochran Q test is also used to assess heterogeneity with a p-value. The final result of the meta-analysis was a forest plot with a pooled odds ratio (OR) value and effect size in each study. The results of the plot funnel were also analyzed for publication bias assessment on the final results of the meta-analysis. The final value used to answer the research objective is the pooled odds ratio value which indicates the combined OR value of several studies. It shows how big the risk factors of each variable studied are. After obtaining the meta-analysis results from the five articles obtained, the authors included the results with the best meta-analysis compared to other variables based on the number of studies and the OR value of each study combined with the appropriate confidence interval.

## RESULT

### Combined Results

#### Individual Study Results

The results of the article selection were obtained from 5 manuscripts that met the criteria. The manuscript reviewer then extracted data on the author’s name, year of publication, type of design, number of samples and estimated sample age, type of hormonal contraceptive therapy used, and statistical results of each study, then tabulated.

**Table 1.** Systematic Review of Electronic-Based Health Promotion Models on the Use of KB IUD Post Placenta

| Study | Settings | Objectives | Study Design | Sample Sizes | Participant Characteristic |
| --- | --- | --- | --- | --- | --- |
| Hersh, 2018 | Colombia | Assessing whether video-based contraceptive | Multicenter randomized, controlled trial | 240 | Structured conversation with a trained counselor or |
|  |  | education can be an efficient addition to contraceptive counseling and achieve the same contraceptive knowledge acquisition as conversation-based counseling. |  |  | A 14-minute video provides the same information. Both groups received written material and were invited to ask questions. Counselor's questions. |
| McCarthy 2019 | Palestine, Bolivia, and Tajikistan | Development and evaluation of contraceptives behavioral interventions delivered by mobile phone to young people in Tajikistan, Palestine, and Bolivia. | Interventions are developed using behavioral science and evaluated by randomized, controlled trials in each country | 1310 | Respondent criteria: age, gender, ethnicity, occupation, level of education, intention at the beginning, the method at baseline, number of children, marital status, and results of admissions at follow-up |
| Tiwari 2019 | India | Data were collected from the mHealth intervention area in India to test the association between exposure to mHealth tools for family planning and behavioral predictors | A post-intervention quasi-experimental study with a 2x2 factorial design | 831 | Men and women from the state of Bihar |
| Stephenson<br>2020 | London | To evaluate interactive websites to assist with informed choices contraception | parallel single-blind experiment | 927 | Ladies between 15 and 30 years of age with present or prospective contraceptive needs, who can read English, have an active email account and an internet connection, and are willing to be tracked for six months. |
| Burapasikarin<br>2020 | Thailand | To evaluate whether postpartum educational videos about MKJP can increase postpartum MKJP use at 6-8 weeks postpartum and to assess why postpartum women do not receive MKJP. | Randomized-controlled trial | 270 | Postpartum women who are > 20 years old and willing to participate are recruited |

Data processing was conducted using Rev Man 5.3 comprehensive meta-analysis software by calculating the effect size, weight, and heterogeneity of effect size to determine the model of combining research and forming the final results of a meta-analysis in the form of forest plot and funnel plot. The following are the results of the meta-analysis and forest plot diagrams, as follows:

The heterogeneity test shows a value ρ greater than 0.05, which is ρ =0.88 with I^2 =^ 0%), meaning that the variation between studies is homogeneous. The Random Effects Model yielded a pooled odds ratio of 1.07 (95& CI 0.26-4.39). The value ρ = 0.93 so that there was no difference between the treatment and control groups, and the data were homogeneous, an odd ratio of 1.07 was obtained. It was found that video use could increase 1.07 times the knowledge, skills, and decision-making of post-placental IUDs compared to those who did not use video. The forest plot shows the information of each study studied and estimates the overall results, visually indicating the magnitude of variation or heterogeneity. After determining the model, the weight of each trial was calculated to obtain the meta-summary analysis’s effect or final results. Based on the forest plot above, there are no meaningful results because it intersects the vertical line and the odds ratio of 1.07, so the data is homogeneous.

The funnel plot is a diagram that explains the possibility of publication bias, showing the relationship between side effects and sample size or standard error of effect size from the various studies studied. Figure 3 presents a funnel plot in this article’s meta-analysis.

**Figure 3.**
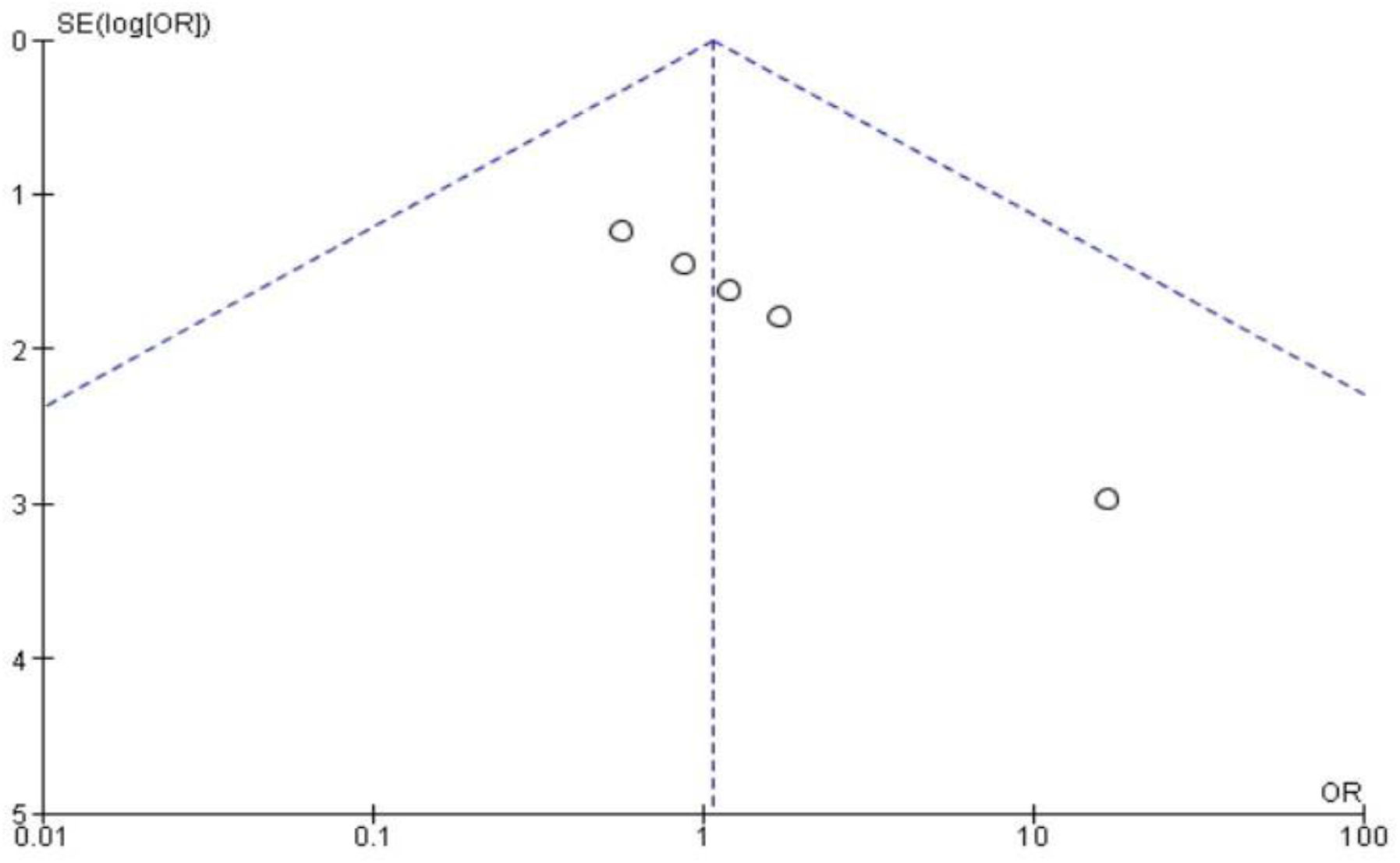
Funnel Plot Diagram

Based on the image indicates that there is a publication bias in the results of this meta-analysis.

## DISCUSSION

About 45% of pregnancies in the UK are unplanned despite the wide range of effective contraceptive methods available for free, and abortion rates in England and Wales have not changed much since 2011. Preventing unwanted pregnancy involves many steps, including timely education, awareness, and socially influenced behaviors to strive for, choose and use contraception consistently and correctly. Healthcare has a crucial role to play by supporting people to choose and use the correct method that best suits their needs, but many people are unaware of the various methods available to them. The contraceptive pill and condoms are well-known and widely used, but they are not the most effective methods of contraception (J. Stephenson, Bailey, Gubijev, D’Souza, Oliver, Blandford, Hunter, Shawe, Rait, Copas, et al., 2020).

Husband/partner support for birth control can affect modern contraception. Socio-demographic factors, couples’ fertility choices, and communication about family planning are known to affect contraceptive use. Perceived husband/partner consent was associated with a threefold chance of using women’s modern contraception and remained significantly associated with a 1.6-fold chance after controlling for contraceptive accessibility and self-efficacy. Husband/partner encouragement was initially significantly associated with contraceptive use and became insignificant after adjusting for socio-demographic factors and couple communication. Perceived husband/partner consent, apart from a woman’s sense of self-efficacy and perceptions of contraceptive accessibility, appears vital and positively associated with the current use of modern contraception. Improved couple communication can help women identify their husband/partner’s approval. The difference between the meaning of consent and encouragement must be explored. Interventions involving information and communication campaigns aimed at men and encouraging their participation in family planning may increase the prevalence of contraception (Prata N, Bell S, Fraser A, Carvalho A, 2015).

Long-acting reversible contraception (LARC), which includes contraceptives, intrauterine systems, implants, and injections, is at least 20 times more effective than oral contraceptive pills and condoms, but these methods are less well known, and not all services have the capacity to customize them. More and more women are turning to online sources of information about sexual health, but information of varying quality and accuracy and misconceptions about contraception are standard. Hormonal contraceptive methods have many potential benefits besides fertility control, including acne treatment, reduced menstrual pain, milder periods or no bleeding, and reduced premenstrual symptoms. However, women may be more aware of contraception’s risks and side effects than the benefits. Interactive digital interventions increase knowledge of contraceptive uptake, more effective contraceptive methods, and contraceptive compliance and reduce unplanned pregnancies. Digital interventions offer the advantage of accuracy and fidelity of intervention content and the potential to reach a broad audience at a relatively low dissemination cost. We, therefore, developed an interactive website to help inform the choice of contraceptive methods and then conducted randomized controlled trials to evaluate their impact on the clinic population (J. Stephenson, Bailey, Gubijev, D’Souza, Oliver, Blandford, Hunter, Shawe, Rait, Brima, et al., 2020).

The puerperium period is vital to promote family planning because puerperium mothers are highly motivated to avoid rapid and recurrent pregnancies. This study shows that providing knowledge about long-term contraceptive methods (MKJP) in postpartum wards using 7-minute animated videos can increase postpartum MKJPC. Although the percentage is also clearly increasing in the non-video group, it is much lower than in the video group (Burapasikarin et al., 2020).

Among the six RCTs assessing contraceptive knowledge, two found evidence of increased knowledge for those receiving electronic interventions (Dewart et al., 2019). A study of women hospitalized for childbirth in Columbia evaluated video contraceptive counseling compared to receiving the same information in a face-to-face conversation with a counselor. The video featured a nurse who provided standard contraceptive information and was about 14 minutes long. The intervention group also had the opportunity to discuss contraceptive options with a counselor, and both groups received written educational materials. No statistically significant differences in knowledge (measured immediately after intervention) were found between the two groups except for knowledge of the definition of family planning, where the effect supported the receipt of information from a counselor (p=0.01 in post-intervention group comparisons). Other measurable knowledge items focus on the efficacy of various methods of contraception (Hersh et al., 2018a).

Electronic platforms provide diverse opportunities for disseminating contraceptive education, although technology must be designed according to the evaluation of content based on health education and behavioral theory. For example, in a study of three video interventions (method-based, motivational, and combination-based), receiving content focused on methods was associated with statistically significant effects related to method choice outcomes and pregnancy incidence. In addition, given that some electronic interventions are conducted similarly to direct counseling methods when followed by the opportunity to speak with a health care provider, the use of electronic media to provide effective contraceptive education can have significant implications related to reducing staff time load (Dewart et al., 2019).

Previous randomized and non-randomized studies assessed the impact of immediate postpartum contraceptive education on postpartum contraceptive use, and the results were inconclusive (Zerden et al., 2015). The 7-minute animated educational video has proven to be a good tool in providing information for postpartum women during their hospital stay. However, about a fraction of women who do not use MKJP admit that they do not know MKJP. Additional qualitative exploration in this group of postpartum women should be helpful. As a result, healthcare providers can implement different education delivery approaches to increase knowledge about the MKJP (Burapasikarin et al., 2020).

In addition, the reasons for not using MKJP shown in this study inform strategies to increase the use of MKJP in the postpartum period. They include MKJP education on prenatal visits. Because many women do not receive prenatal care at the facility where they gave birth, decisions about family planning before delivery are not easily communicated to hospital staff. Given staff limitations, providing individualized contraceptive counseling is a challenge. Furthermore, contraceptive counseling is not currently incorporated into routine prenatal care. In this study, we sought to show that video-based contraceptive education can be an efficient addition to counseling and achieve the same level of family planning knowledge as conversation-based counseling (Hersh et al., 2018b).

Interactive digital interventions are effective at increasing the absorption of contraceptive knowledge and helping to choose more effective methods of contraception and contraceptive compliance, as well as reducing unplanned pregnancies. Digital interventions offer the advantage of accuracy and fidelity of intervention content and the potential in order to reach a broad audience at a minimal cost of deployment (McCarthy, 2019; J. Stephenson, Bailey, Gubijev, D’Souza, Oliver, Blandford, Hunter, Shawe, Rait, Brima, et al., 2020). Mobile phones serve as a tool that can improve women’s participation in their health care and their families. Mobile phone-based health tools that are easy to use have opened up new opportunities to provide women with information that is important for their health and well-being, one of which is information about birth control. Thus, technology-based communication tools can help initiate dialogue at the community level that can lead to the dissemination of information, as well as negotiation and mediation of the use of birth control (Tiwari, 2019). Videos about birth control can be inserted into mobile phones to facilitate access to information for women, couples, and families that impact the increased use of contraception, especially postpartum IUDs.

## CONCLUSION

Health promotion models that use electronic-based tools (video) 1.07 times can improve knowledge, skills, and decision-making using long-term contraceptives, especially IUDs, compared to those that do not use video

## Data Availability

All data produced in the present work are contained in the manuscript

